# Acute resistance exercise-induced changes in IL-6, IL-10, and IL-1ra in healthy adults: a systematic review and meta-analysis

**DOI:** 10.1101/2023.05.10.23289790

**Authors:** Miriam Ringleb, Florian Javelle, Simon Haunhorst, Wilhelm Bloch, Lena Fennen, Sabine Baumgart, Sebastian Drube, Philipp A. Reuken, Mathias W. Pletz, Heiko Wagner, Holger H. W. Gabriel, Christian Puta

## Abstract

**Background:** Myokines, released from the contracting muscle, enable communication between the working muscles and other tissue. Their release during physical exercise is assumed to depend on mode, duration, and intensity.

**Objective:** The aim of this meta-analysis is to examine the acute changes in circulating levels of the myokines IL-6, IL-10, and IL-1ra induced by a bout of resistance exercise and to consider potential moderators of the results.

**Methods:** Systematic literature search was conducted for resistance exercise intervention studies measuring IL-6, IL-10, or IL-1ra before and immediately after resistance exercise in healthy individuals. Random effects meta-analysis was performed for each myokine.

**Results:** A small to moderate positive effect of resistance exercise for IL-6 and a moderate to large positive effect for IL-1ra were detected. For IL-10 no significant effect was observed. No moderators (training status, type of exercise, risk of bias, age, exercise volume, exercise intensity, exercise dose) of the results were detected.

**Conclusion:** This systematic review and meta-analysis clearly showed the immediate positive effects of an acute resistance exercise session on IL-6 and IL-1ra levels.

## 1 Introduction

Cytokines are biochemical mediators typically secreted by immune cells known to exert pleiotropic effects such as communication and coordination between various target cells in the human body (Ouchi *et al*. 2011). They can act in an autocrine, paracrine, or endocrine manner to induce or suppress their synthesis and regulate the production of other cytokines and their receptors (Chauhan *et al*. 2021; Fiuza-Luces *et al*. 2018). These low molecular weight proteins are involved in multiple processes, including immune regulation, inflammatory reactions, and the maturation of blood cells (Petersen & Pedersen 2005). Based on the secreting cell or mechanism of action, cytokines have been broadly categorized as chemokines (synthesized to induce leukocyte migration), interleukins (synthesized by leukocytes), lymphokines (synthesized by lymphocytes), adipokines (synthesized by adipocytes), and monokines (synthesized by monocytes and macrophages) (Chauhan *et al*. 2021; Ouchi *et al*. 2011). In the past two decades, multiple studies have shown that muscle cells can also produce a large variety of cytokines (Chauhan *et al*. 2021; Fiuza-Luces *et al*. 2018; Pedersen 2011b). They are now referred to as myokines (Pedersen *et al*. 2007; Puta & Gabriel 2020; Roth *et al*. 2018).

Myokines play a major role in human metabolism via their involvement in central homeostatic mechanisms like lipolysis in skeletal muscles, fat oxidation in adipose tissue, and their contribution to metabolic homeostasis (Fiuza-Luces *et al*. 2018). By communicating between the muscle and other organs, including the brain, bone, and vascular system, myokines affect human cognition, mental health, bone formation, and endothelial cell function (Severinsen & Pedersen 2020). Furthermore, it is well-established that myokines greatly impact the regulation of immunological processes in response to physical activity (Pedersen 2011b; Pedersen & Febbraio 2012). Released by concentric muscle contractions (Pedersen *et al*. 2007), they mediate the health-promoting effects of exercise and contribute to the protection against diseases associated with low-grade inflammation, such as cardiovascular diseases, type 2 diabetes, and mental health disorders (Mucher *et al*. 2021; Pedersen *et al*. 2003; Pedersen & Febbraio 2008).

However, myokines are not only released by muscle fibers, but in the regenerating muscle also from infiltrating neutrophils and macrophages (Ulven *et al*. 2015), from fibro-adipogenic progenitors (Islam *et al*. 2021), and from satellite cells (Joisten, Schumann *et al*. 2020; Kakanis *et al*. 2014). Nevertheless, growing evidence from biopsy, gene sequencing, or blood collection studies (Roth *et al*. 2018; Schild *et al*. 2016; Steensberg *et al*. 2003) suggests that the release of IL-6 from working muscle myocytes is far more important than that from immune cells for its systemic increase.

The first cytokine identified as a myokine was the muscle-derived interleukin (IL)-6 in 2000 by Pedersen’s laboratory (Steensberg *et al*. 2000). Initially, IL-6 was known as a pro-inflammatory cytokine (Pedersen & Hoffman-Goetz 2000). Acute trauma or infection causes a local and systemic rise in IL-6, leading to the activation and release of hepatocyte-derived acute phase proteins (APP), such as C-reactive protein (CRP) (Petersen & Pedersen 2005; Tanaka *et al*. 2014). This rise in IL-6, APP, and several other cytokines ensures a rapid and targeted immune response (Petersen & Pedersen 2005).

However, current literature shows that an increase in muscle-derived IL-6 following exercise exerts anti-inflammatory effects without causing a rise in pro-inflammatory TNF-α or CRP. Beyond that, as muscle-derived IL-6 release, paradoxically, even led to decreasing levels of TNF (Fiuza-Luces *et al*. 2018; Starkie *et al*. 2003) and IL-1 (Petersen & Pedersen 2005), it has been postulated that anti-inflammatory processes are initiated via myokine release. Moreover, IL-6 release has been shown to stimulate the production of the anti-inflammatory cytokines IL-1ra and IL-10 (Fiuza-Luces *et al*. 2018; Steensberg *et al*. 2003), which are also considered myokines. Finally, IL-6 and IL-10 release contributes to an anti-inflammatory environment by inhibiting Th1 cell activity and promoting Th2 cell function, which promotes cellular as well as humoral immunity and inflammatory response (do Brito Valente *et al*. 2021; Puta & Gabriel 2020).

Existing literature suggests that acute and chronic physical activity are associated with changes in myokine concentrations and their mRNA expression depending on exercise duration, mode, and intensity (Gebhardt & Krüger 2022; Pedersen *et al*. 2007). Although chronically elevated cytokine concentrations are associated with disease states and inflammation (Pedersen & Hoffman-Goetz 2000), this cannot be demonstrated for acutely elevated myokine concentrations after exercise. These acute positive effects have been shown for both endurance and resistance exercise. For instance, Schild et al. (2016) showed that 50 minutes of cycling led to significantly increased IL-6 and IL-10 levels. Interestingly, some authors like Fischer et al. (2004) have detected stronger effects after three hours of dynamic two-legged knee-extensor exercise at 50% of the individual maximal power output. This led to a 16-fold increase in IL-6 mRNA and a 20-fold increase in plasma IL-6. Significant changes after an acute bout of exercise were also observed for IL-1ra and IL-10 (Pekkala *et al*. 2013; Schild *et al*. 2016). While there is an extensive body of literature investigating continuous endurance exercises (Broholm *et al*. 2008; Chan *et al*. 2004; Fischer *et al*. 2004; Hiscock *et al*. 2004; Huh *et al*. 2014; Keller *et al*. 2005; Mucci *et al*. 2000; Nieman *et al*. 2001; Nieman *et al*. 2002; Nieman *et al*. 2003; Ostrowski *et al*. 1998; Pekkala *et al*. 2013; Schild *et al*. 2016; Windsor *et al*. 2018), the general cytokine response to intermittent exercise modes has only been scarcely evaluated, and determinants of the response remain comparably poorly studied. This is especially true for resistance exercise, albeit it represents the most direct form of volitional muscle fiber recruitment and an easily implementable exercise form. Yet, even though most findings on the effects of resistance exercise on myokine levels mirror those made in the context of endurance exercise, their magnitudes greatly vary between studies, with some even reporting no changes (Cornish *et al*. 2018; Pereira *et al*. 2013; Windsor *et al*. 2018) or a reduction (Lira *et al*. 2020) in myokine concentrations.

To better understand the myokine response to exercise, this systematic review and meta-analysis aims to examine and quantify the acute changes in circulating levels of the myokines IL-6, IL-10, and IL-1ra induced by a bout of resistance exercise and to characterize how different exercise parameters influence this response. As indicated, IL-6 is the best-known and most studied myokine, and therefore it is of great interest to know whether there is a consistently significant effect of resistance exercise in the studies investigating these effects. Furthermore, since the release of IL-6 has been shown to induce an increase in IL-10 and IL-1ra (Fiuza-Luces *et al*. 2018; Steensberg *et al*. 2003), the next logical step is to also investigate their release systematically after resistance exercise.

In line with current knowledge, we hypothesized that acute resistance exercise would increase IL-6, IL-10, and IL-1ra levels in healthy adults. Furthermore, we hypothesized that the dose of exercise would be positively associated with myokine release. The other moderators were considered in an explorative manner. Fostering the understanding of the relationship between exercise modalities and myokine response would, for example, help to predict training outcomes and to optimize exercise recommendations for populations with different requirements, such as patients with cancer, multiple sclerosis, or post-viral infection syndromes.

## 2 Methods

The Preferred Reporting Items for Systematic Reviews and Meta-Analyses (PRISMA) 2020 guidelines (Page *et al*. 2021) were adopted for the literature search and writing process. The PRISMA checklist is provided as a supplementary file. The protocol was pre-registered on PROSPERO (Registration number: CRD42022327039, last amendment date: 09/05/22). Three different meta-analyses were performed to evaluate blood concentration changes from pre- to immediately post-exercise intervention of 1) IL-6, 2) IL-10, and 3) IL-1ra.

### 2.1 Literature search

The literature search was conducted using databases MEDLINE (via PubMed), Web of Science, Cochrane Library, and SportDiscus from 20/01/22 to 18/02/22. The search query was created based on MeSH terms and related vocabulary covering the main domains of resistance exercise and interleukins (see Table 1). The exact search syntax for each database can be accessed via PROSPERO. Additionally, the reference lists of the included studies were screened.

**Table 1.**
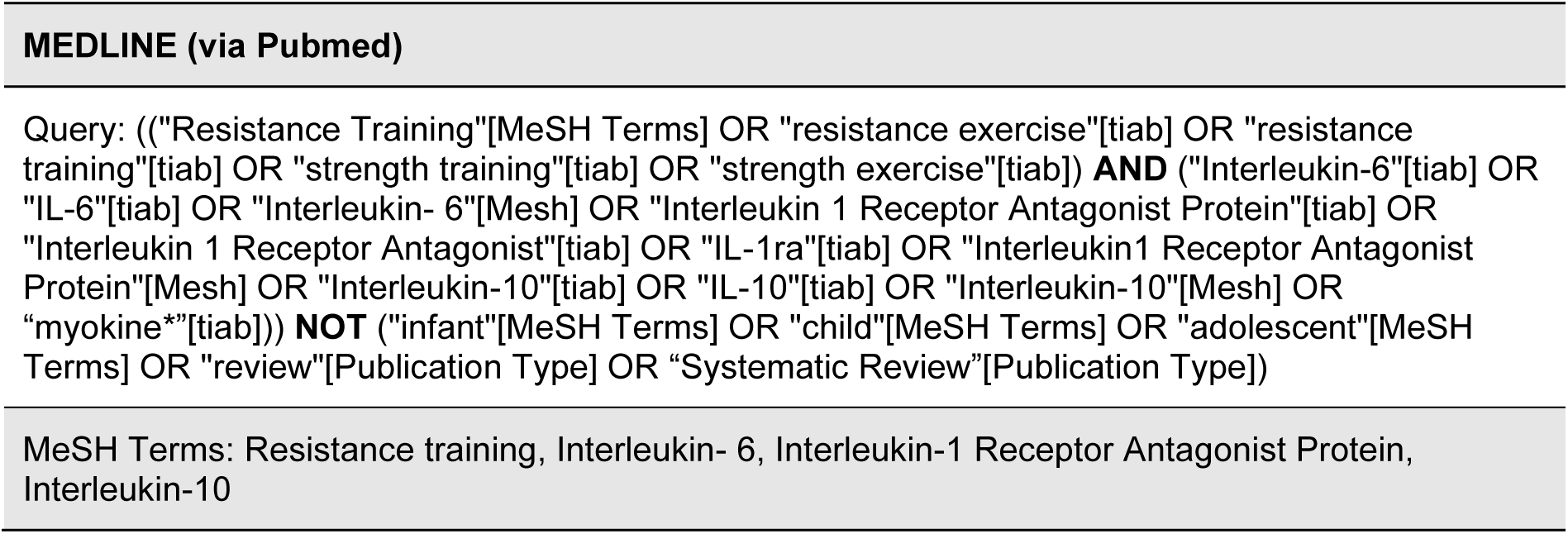
*MeSH terms and search query*

### 2.2 Eligibility criteria

Eligibility criteria were determined using a PICOS (participants, interventions, comparison, outcomes, study design) approach.

1. Participants: Healthy participants between 18 and 60 years were included in the review. Studies examining participants older than 60 years were excluded. Individuals suffering from any disease or injury (chronic or acute) were excluded from the review as well.
2. Interventions: All studies comprising a single resistance exercise session defined as concentric or eccentric muscle actions overcoming externally applied resistance were included in the review. If a study included an exercise program, but an acute measurement was conducted before and after a single resistance training session, this acute intervention was included in this review. Studies combining resistance training with endurance exercise, conducting an exercise program over several weeks, or combining exercise with additional treatments possibly altering the physiological response to exercise were excluded from the review. In addition, studies including interventions such as yoga, stretching, breathing exercises, or other types of exercise that do not fit the definition of resistance exercise were excluded from the review. When studies had multiple interventions, only the intervention groups assessing the effect of resistance exercise were included in the review.
3. Comparison: To be considered, the study’s outcome parameters must have been measured pre-exercise and immediately post-exercise. Studies with no baseline or follow-up measurements later than five minutes post-exercise were excluded from the review.
4. Outcomes: Studies assessing the changes in blood serum or plasma concentration of either IL-6, IL-10, or IL-1ra were included in the review.
5. Study design: Randomized and non-randomized clinical trials published in English or German in a peer-reviewed journal. Case studies, animal studies, reviews, cross-sectional or retrospective studies, and longitudinal study designs were excluded from the review.

### 2.3 Study selection

The studies found in the literature databases were uploaded to Rayyan (https://www.rayyan.ai/), a free platform allowing the authors to screen the records independently and blinded to the decision of the other. MR and SH made the study selection. First, duplicates were removed. Thereafter, titles and abstracts were screened, and studies not fitting the eligibility criteria were excluded. Disagreements between the reviewers after either the screening of titles and abstracts or full-text screening were solved by discussion. Figure 1 outlines the selection process.

**Figure 1.**
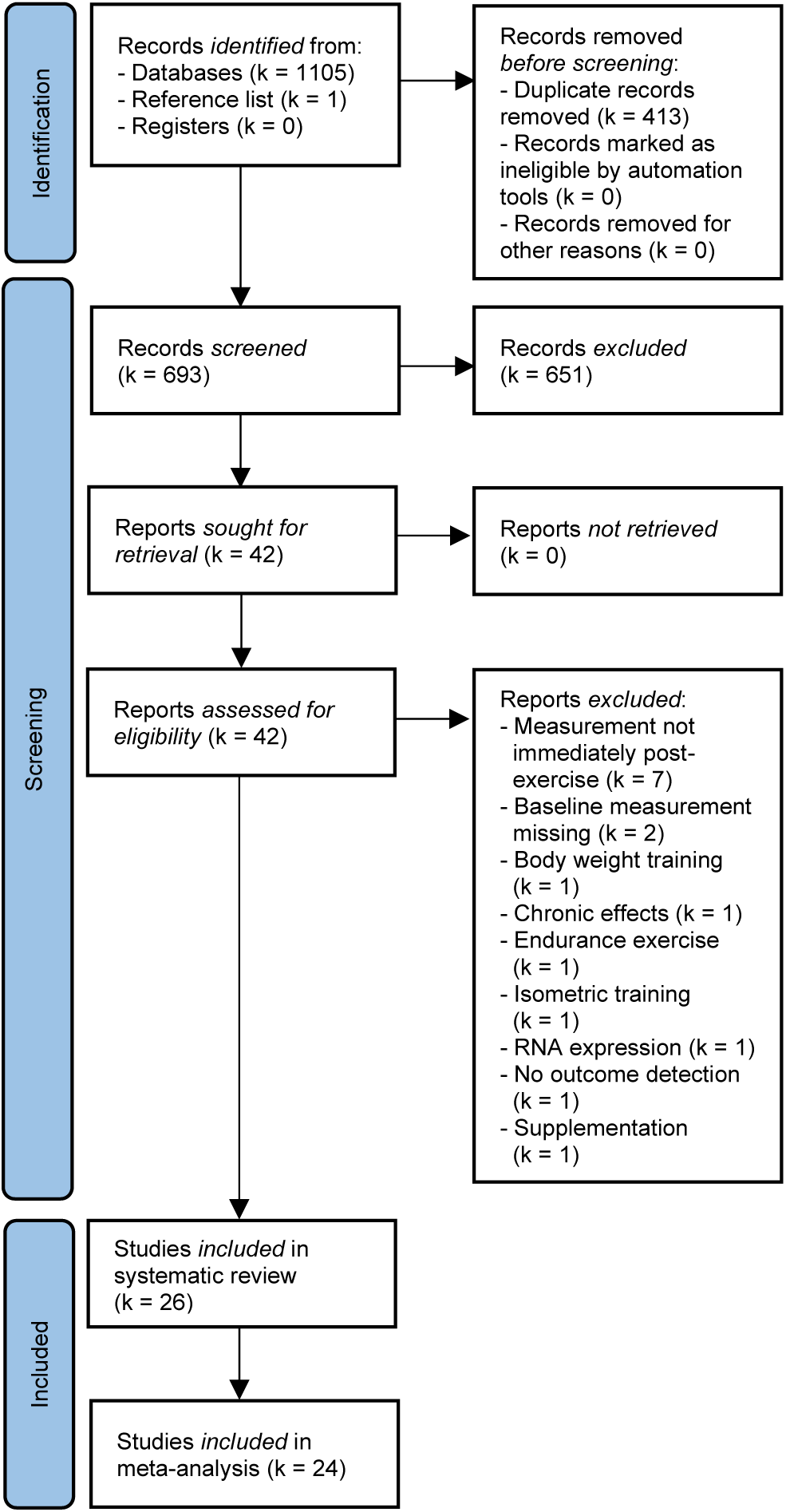
Flowchart of literature search and study selection

### 2.4 Data extraction

Data extraction was conducted by MR. First, general information like authors, study design, and sample size were extracted from the studies. Second, for each intervention group, myokine levels (pre- and immediately post-intervention) with mean and standard deviation (or standard error of the mean or 95% confidence interval) were extracted. Third, participant characteristics such as sex, age, height, weight, and body mass index (BMI) were extracted. Fourth, the resistance training status was also collected. Participants were considered inexperienced if they were described as untrained or sedentary or did not participate in any kind of regular resistance training six months before testing. Moreover, the resistance exercises, training parameters (number of sets and repetitions, intra- and inter-set rest), and intensity were extracted. Furthermore, we computed the dose as the product of intensity and volume, as adopted by Herold et al. (2019). The exercise intensity is mostly provided as one-repetition maximum (1RM) or multiple-repetition maximum (MRM). Since only the 1RM is considered for calculating the dose, the MRM is, based on Baechle & Earle (2008), converted to the 1RM if this is the only one reported in the studies. Finally, the time of the day and the type of sample gathered were recorded.

If the authors did not provide exact values, the WebPlotDigitizer digitization program (https://automeris.io/WebPlotDigitizer/) was used to extract plotted data. In parallel, corresponding authors were contacted to retrieve missing data.

### 2.5 Risk of bias assessment

The risk of bias in individual studies was assessed using the ROBINS-1 tool (Sterne *et al*. 2019). As it evaluates the quality of studies performing a controlled pre- and post-study intervention, this tool is the most suitable quality assessment for this review. The ROBINS-1 tool consists of seven domains appraising the risk of bias. In the case of this review, the domain “bias due to deviations from intended interventions” was not considered since a comparison with a control group is not necessary. Every domain was evaluated via signal questions that were derived from Palmowski et al. (2020). Based on this, the individual domains were classified as having either a low, moderate, serious, or critical risk of bias or no information available on which the judgment could be based. Overall, a study was judged to be at low risk of bias if all domains were assessed to be at low risk of bias and moderate risk when at least one domain shows a moderate risk of bias. A serious risk of bias was present when the study was ranked at serious risk in at least one domain but not at critical risk in any domain. With at least one domain at critical risk of bias, the overall study was judged to be at critical risk of bias. No judgment was possible if there was no clear indication that the study was at serious or critical risk of bias and a lack of information was found in one or more key domains. The assessment of the risk of bias was done independently by MR and SH. The inter-rater coefficient of correlation of the overall judgment of the risk of bias was r=0.81. Any disagreements were then solved by discussion.

### 2.6 Effect Sizes

Changes in myokine concentrations were converted to standardized mean differences, calculated as Hedges’ g (g) to account for small study sample sizes. All studies had within-subject designs with pre- and post-intervention values. All effects had the same direction, and a positive effect size denotes an increase in those three myokines after the exercise intervention. By convention, Hedges’ g values of 0.2, 0.5, and 0.8 are respectively considered small, moderate, and large.

The Hedges’ g (SMD=MD/SDpooled) and its standard error were computed according to the recommendations from Borenstein et al. (2009) (4.15 – page 24) and the Cochrane Handbook (chapter 23, section 23.2.7.2). The standard error was computed using the imputation of a correlation coefficient at 0.7 (chosen based on literature and available data) between the immune values pre- and post-intervention. To assess the reliability of this coefficient, sensitivity analyses were run ±0.15 the chosen coefficient (between 0.55 and 0.85 - 0.05 per interval). No differences in the significance of the overall effect, asymmetry, and moderator analyses were identified, underlining the reliability of our results.

### 2.7 Statistical Analysis

All analyses were conducted using R Studio (v1.4.1106), using the packages meta, metafor, and metaviz (Viechtbauer & Cheung 2010). The full R script and all CSV files used for analyses (including the ones of the sensitivity analysis) are available on the Open Science Framework (doi: 10.17605/OSF.IO/5JKFA). Effect sizes across studies were pooled using a random-effect model (Higgins *et al*. 2019). Separate meta-analyses were run for each myokine of interest (i.e., IL-6, IL-10, IL-1ra). According to Viechtbauer and Cheung (2010), influential outliers were estimated using studentized deleted residuals and DFBETAS. An outcome was considered as being an outlier if its studentized residual was greater than three in absolute magnitude. It called for closer inspection if its studentized residual was greater than 1.96 in absolute magnitude. To evaluate if these outliers should also be considered influential cases, the DFBETAS was computed. According to Viechtbauer and Cheung (2010), DFBETAS greater than one is often considered as being an influential case when considering small to medium datasets.

### 2.8 Heterogeneity

The between-study heterogeneity was measured using tau^2^ (variance of the true effect), using Hedges’ estimator (Hedges & Olkin 1985), and further assessed using the I^2^ statistic (measures the percentage of the observed variance reflecting the variance of the true effect rather than sampling error) (Borenstein 2019; Thompson-Lake *et al*. 2017). The prediction interval (PI) was also computed to consider the potential effect of an exercise bout on myokine levels when applied within an individual study setting, as this may differ from the average effect (Riley *et al*. 2011). The Hartung and Knapp method was used to adjust confidence intervals and test statistics (Hartung & Knapp 2001a, 2001b; IntHout *et al*. 2014).

Moderator analyses were planned for age, sex, training status, exercise type, exercise volume, exercise intensity, exercise dose, time of the day, and risk of bias estimated. Subgroup analyses were performed for categorical moderators (i.e., sex, training status, exercise type, time of the day, and risk of bias estimated), and meta-regressions were performed for continuous moderators (i.e., age, exercise volume, intensity, and dose). Multivariate moderator analyses were performed to assess the combined effects of different and non-overlapping exercise modalities (i.e., only exercise dose was used among intensity, volume, and dose) using the metafor package (rma function).

### 2.9 Small study effects

Small study effects (an indicator of potential publication bias) were first assessed by visual inspection of the funnel plots and further assessed using Egger’s test (significant when *p*<0.1, one-tailed). If evidence of asymmetry was found, the trim-and-fill procedure was used to estimate the number of potentially missing effects and to provide an adjusted Hedges’ g estimate (Duval & Tweedie 2000; Egger *et al*. 1997; Viechtbauer & Cheung 2010).

## 3 Results

### 3.1 Study selection

In total, 1,106 studies were found through a systematic literature search in the four databases: Pubmed (449), Web of Science (385), Cochrane Library (212), and SportDiscus (59), and one further study was identified through the screening of reference lists. After removing duplicates, 693 titles and abstracts were screened for eligibility. As a result, 42 reports were sought for retrieval and were assessed for eligibility. A total of 26 studies were included in the systematic review, and 24 were included in the meta-analyses (missing values in two studies - Figure 1).

### 3.2 Study characteristics

Twenty-six studies, including 52 intervention groups with a total of 373 (88% male/12% female) participants, were included in this systematic review, among which four intervention groups were not considered for the meta-analysis due to inaccessible data (Barquilha *et al*. 2018; Gadruni *et al*. 2015; Heavens *et al*. 2014). From the total of 24 studies included in the meta-analyses, all 24 investigated IL-6 with 48 effect sizes (Agostinete *et al*. 2016; Benini, Prado Nunes, Orsatti, Barcelos, Orsatti 2015; Brenner *et al*. 1999; Cui *et al*. 2017; Fatouros *et al*. 2010; Gadruni *et al*. 2015; Gerosa-Neto *et al*. 2016; Gordon *et al*. 2017; Goto *et al*. 2020; Ihalainen *et al*. 2014; Ihalainen *et al*. 2017; Izquierdo *et al*. 2009; Joisten, Kummerhoff *et al*. 2020; Krüger *et al*. 2011; Lira *et al*. 2020; Luk *et al*. 2021; Marcucci-Barbosa *et al*. 2020; Nakajima *et al*. 2010; Oliver *et al*. 2016; Phillips *et al*. 2010; Pledge *et al*. 2011; Quiles *et al*. 2020; Rossi *et al*. 2016; Vincent *et al*. 2014), ten examined IL-10 with 22 effect sizes (Agostinete *et al*. 2016; Benini, Prado Nunes, Orsatti, Barcelos, Orsatti 2015; Brenner *et al*. 1999; Cui *et al*. 2017; Fatouros *et al*. 2010; Gerosa-Neto *et al*. 2016; Izquierdo *et al*. 2009; Lira *et al*. 2020; Luk *et al*. 2021; Marcucci-Barbosa *et al*. 2020) and further four analyzed IL-1ra with 12 effect sizes (Agostinete *et al*. 2016; Ihalainen *et al*. 2014; Ihalainen *et al*. 2017; Izquierdo *et al*. 2009). As displayed in Table 2, only one study investigated the effects of resistance exercise on female participants (Luk *et al*. 2021), and two studies had at least one female intervention group investing sex-dependent differences in results (Benini, Prado Nunes, Orsatti, Barcelos, Orsatti 2015; Heavens *et al*. 2014). Two studies did not report their participants’ sex (Barquilha *et al*. 2018; Vincent *et al*. 2014). Participants were, on average, 25.8 (18-51) years old and had a mean BMI of 24.68 kg/m^2^. Sixty percent of the participants were described as experienced in resistance training, whereas 40% had no resistance training experience. Regarding the single exercise session, 48% of the participants underwent a full-body exercise session, whereas 42% of the interventions comprised a lower-body workout and 10% an upper-body resistance training. Session volume (sets x repetitions) ranged from 15 to 345 repetitions, and exercise intensity from 40 to 100% of the 1RM. More than half of the studies did not report their testing time (16 out of 48 intervention groups), yet amongst the reported ones, the majority of the sessions were conducted in the morning. Sixty-nine percent of the studies used enzyme-linked immunosorbent assays (ELISA) to analyze their blood samples. All study characteristics are displayed in detail in Table 2.

**Table 2.**
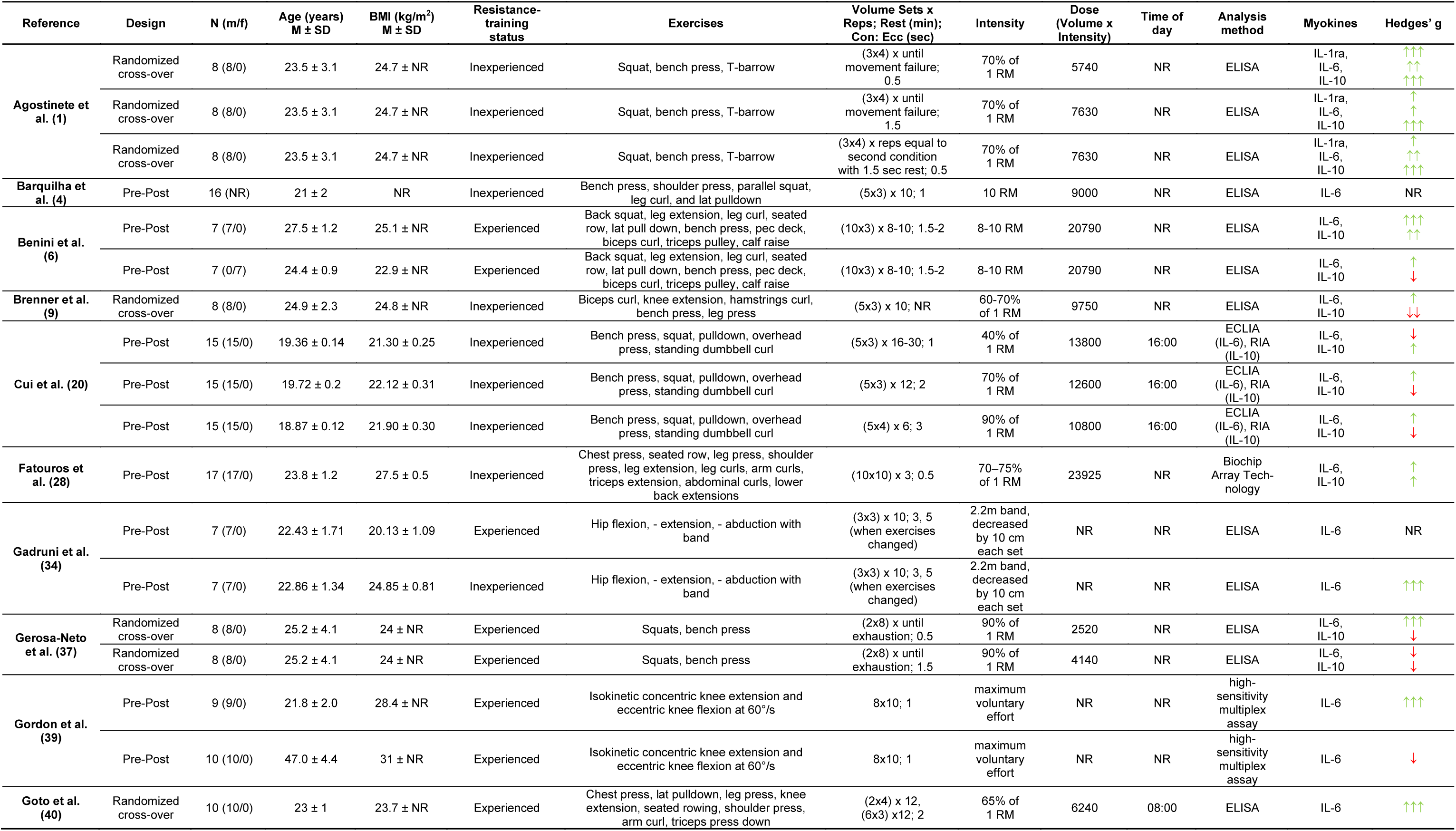
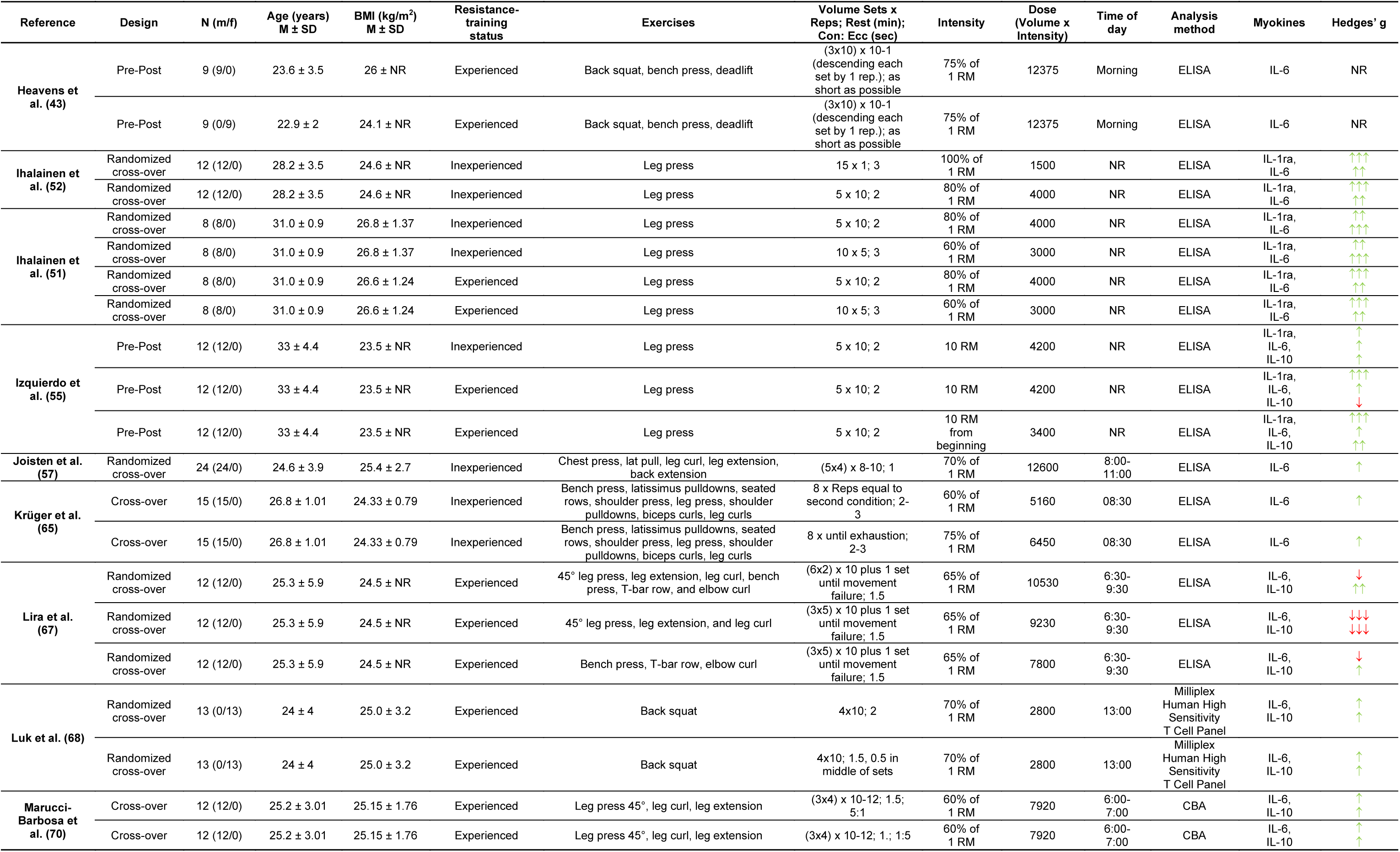
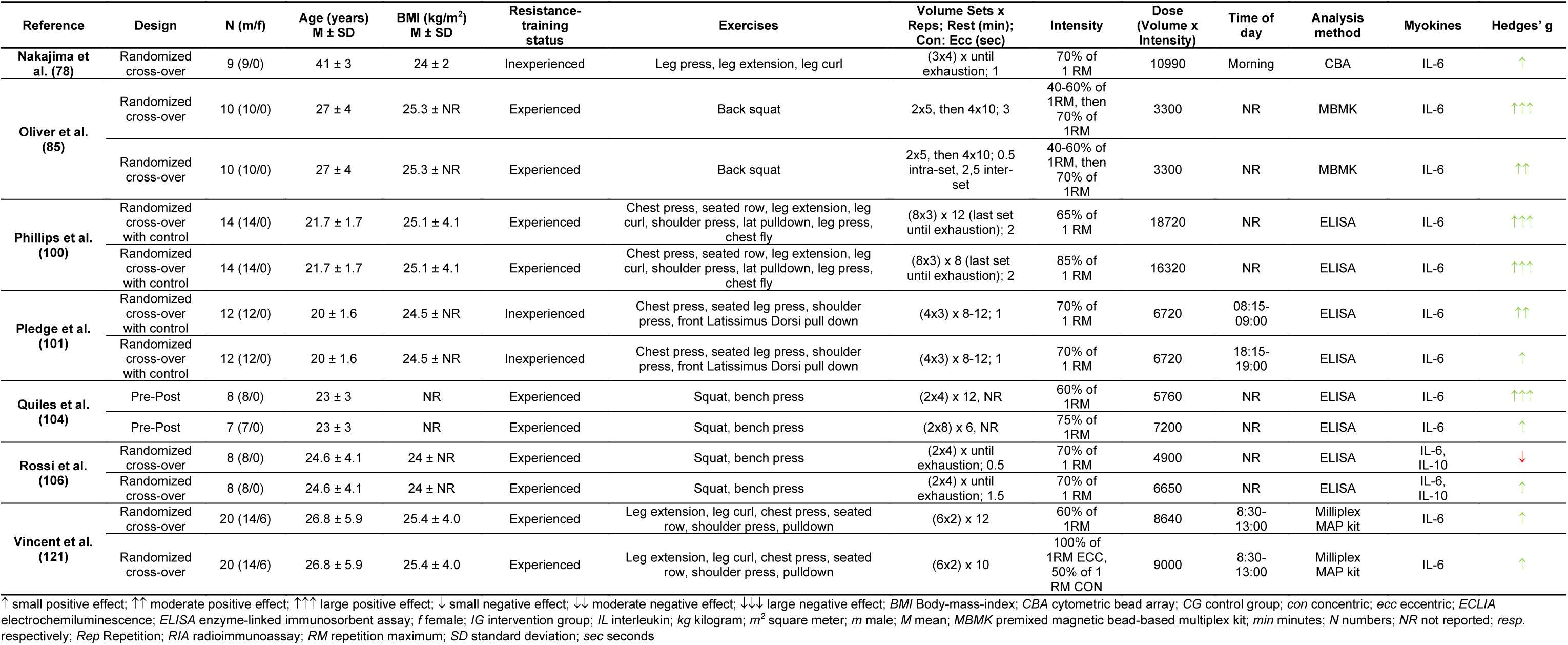
*Study characteristics*

### 3.3 Risk of bias assessment

Only one study was deemed to be at low risk of bias (3%), whereas nine were labeled as moderate (35%). Eight studies were assessed to be at serious risk of bias (31%), and another eight showed a critical risk of bias (31%). No study showed serious or critical risk in the three domains “bias due to missing data”, “bias in the measurement of the outcome”, and “bias in the selection of the reported results”. If one study was considered to be at serious or critical risk of bias, most inconsistencies occurred in the first, the confounding domain. Less than half of the studies showed a low or moderate risk of bias (42%) in this domain. The main reasons for the poor ratings in this domain were the lack of control of nutritional intake, no indication of 24 to 48 hours of rest before the intervention, or differing warm-ups between the participants. Detailed results are shown in Table 3. As more than half of the studies demonstrated a serious to critical risk of bias, the overall quality of the existing literature can be depicted as low.

**Table 3.**
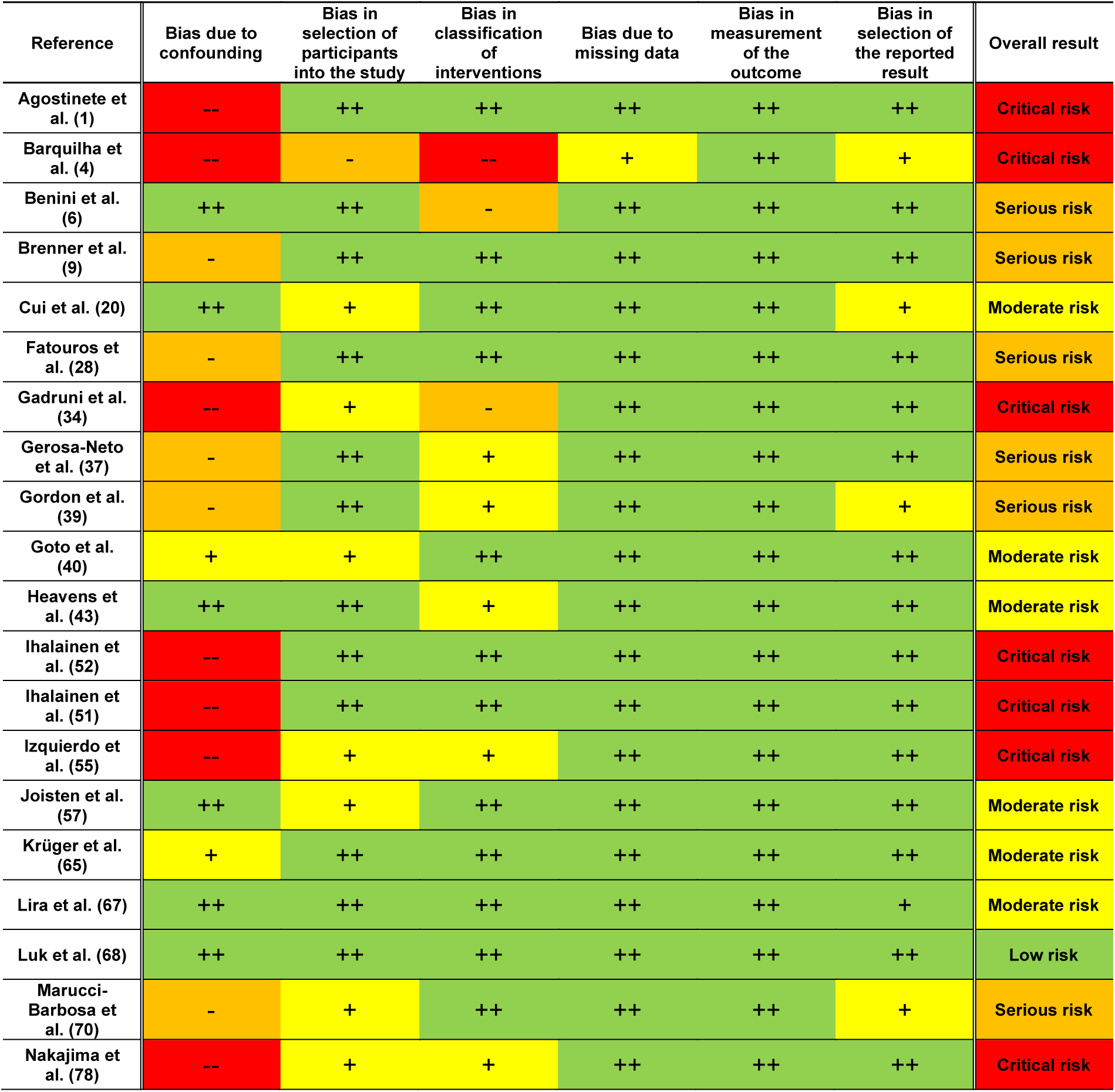
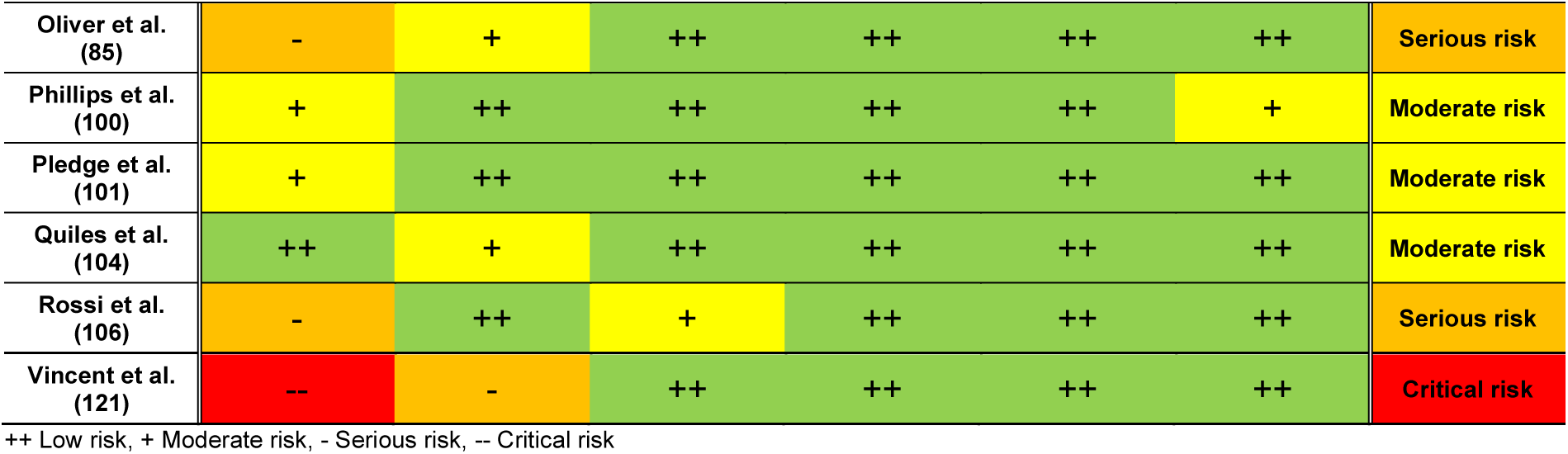
*Risk of bias assessment*

The results of the three meta-analyses performed respectively for IL-6, IL-10, and IL-1ra are presented below and summarized in Table 4.

**Table 4.** *Results Meta-Analyses*

|  | IL-6 | IL-10 | IL-1ra |
| --- | --- | --- | --- |
| g [CI 95%] | 0.39 [0.24; 0.53]** | 0.16 [-0.09; 0.40] | 0.78 [0.55; 1.02]** |
| PI | [-0.61; 1.38] | [-0.96; 1.28] | [0.14; 1.42] |
| I<sup>2</sup> | 71.5% | 75.7% | 44.0% |
| k | 48 | 22 | 12 |
| Participants | 325 (164<sup>a</sup>) | 157 (73<sup>a</sup>) | 40 (40<sup>a</sup>) |
| Tested moderators | Training status, type of exercise, risk of bias, age, exercise volume, exercise intensity, exercise dose | Training status, type of exercise, risk of bias, age, exercise volume, exercise intensity, exercise dose | Training status, age, exercise dose |

\*\* Significance < 0.01, <sup>a</sup> participants counting for at least two effect sizes, CI Confidence interval, g Hedges' g, IL Interleukin, k number of studies included, PI prediction interval

### 3.4 Interleukin-6

#### 3.4.1 Main analysis

The average effect size for IL-6 was small to moderate (k=48, g=0.39, *p*<.001), and its confidence interval was between 0.24 and 0.53, which indicates that in a universe of comparable studies to the one included in this analysis, the mean effect size could fall anywhere in between small and moderate (Figure 2A). This range did not include 0, which revealed that the true effect is very likely to be positive. The heterogeneity was moderate to large (PI [-0.61; 1.38], tau^2^=0.24), with a large part representing the variance of the true effect (I^2^=71.5%). According to the PI, in the universe of populations represented by the included studies, the true effect in 95% of cases will fall between moderate negative and very large positive effects. No outliers were detected. There was a small visual asymmetry (Figure 3A), confirmed by Egger’s test (intercept=-0.63, *p*<.001), but no adjustments were necessary according to the trim-and-fill analysis (SE=4.04).

**Figure 2.**
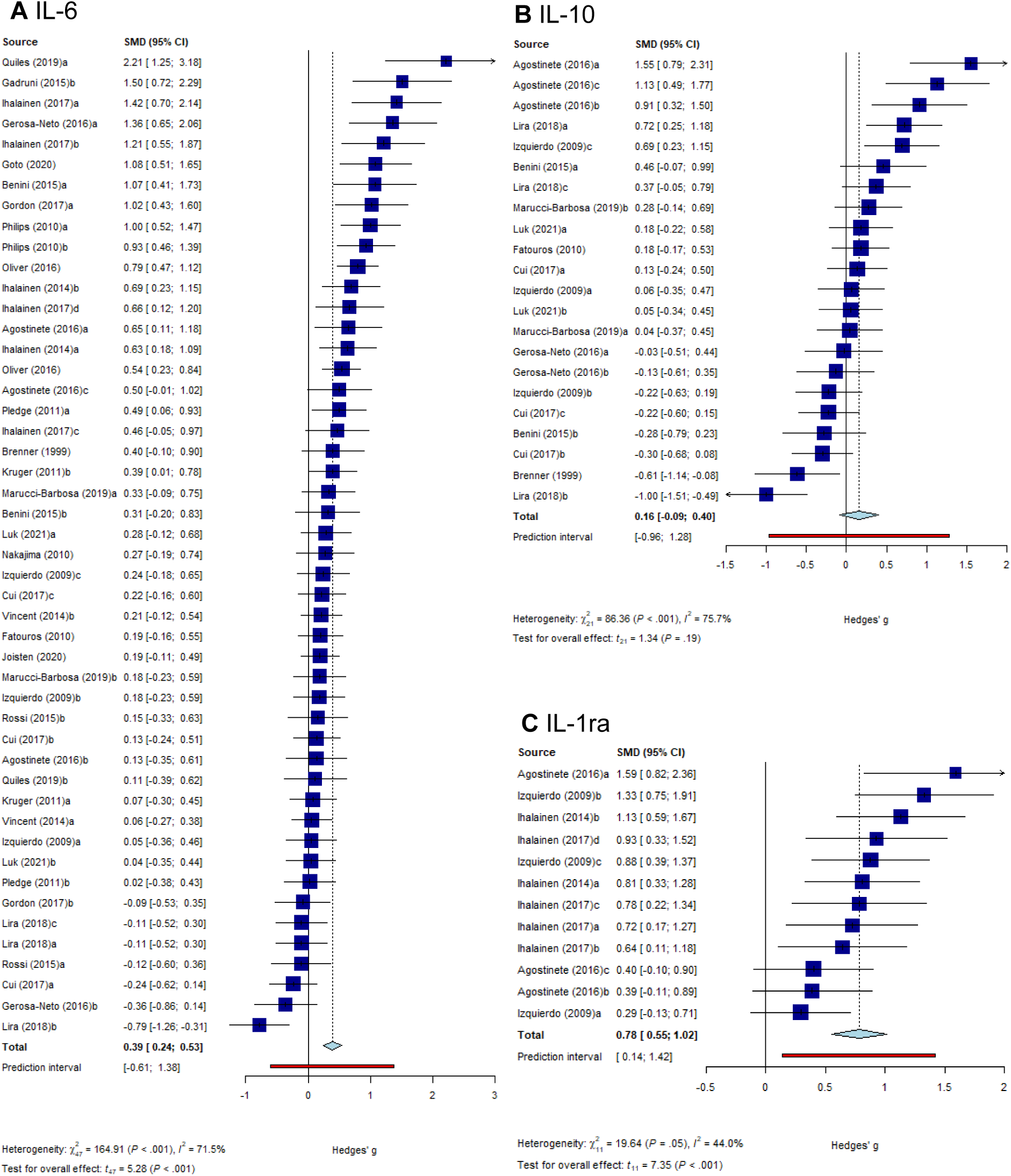
Forestplot for the comparison pre- to post-exercise in IL-6 (A), IL-10 (B), and IL-1ra (C). a-d, correspond to intervention groups/protocols within one study; CI, confidence interval; SMD, standardized mean difference

**Figure 3.**
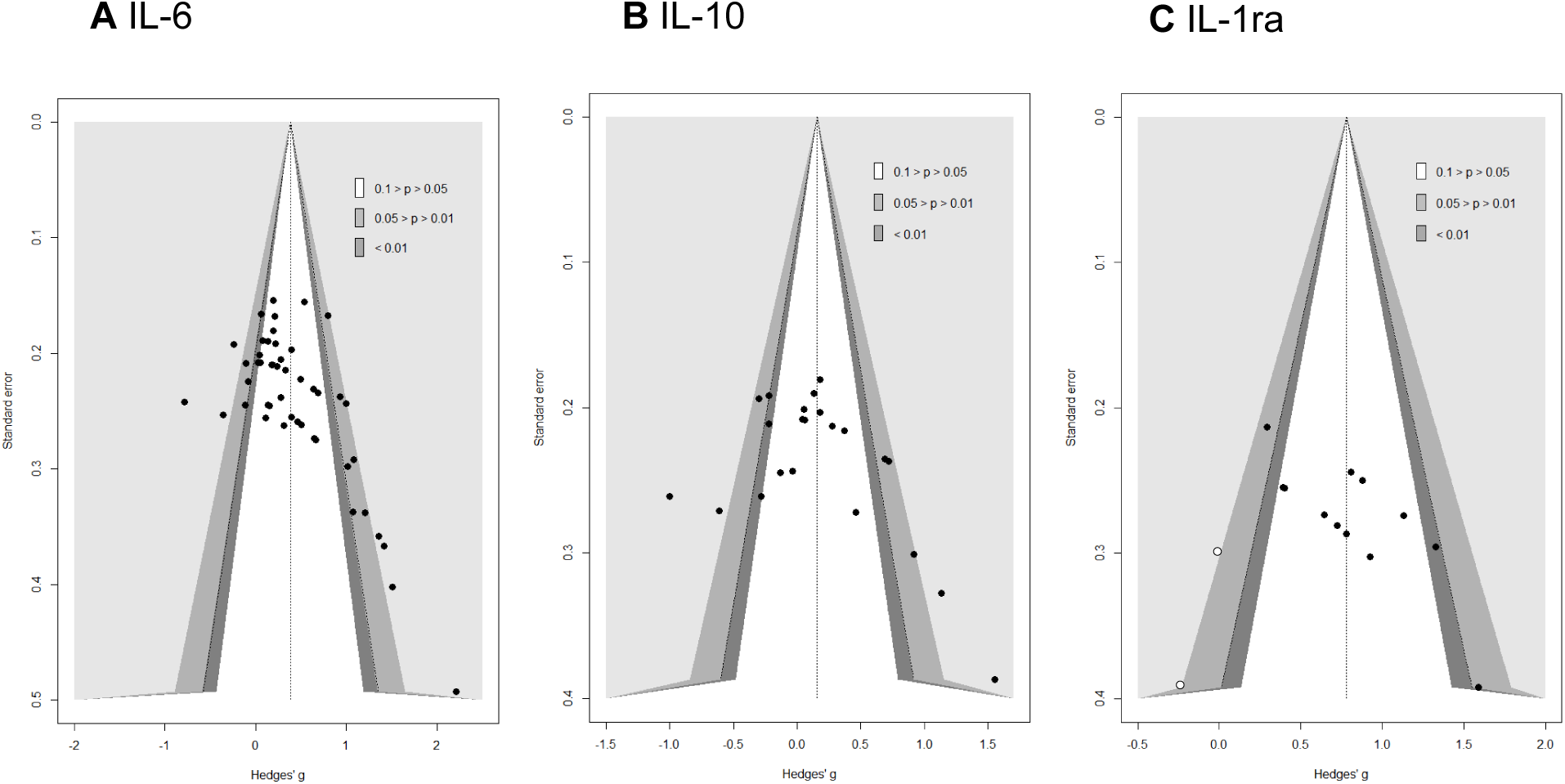
Funnelplot for IL-6 (A), IL-10 (B), and IL-1ra (C)

Due to the skewed distribution in sex (4 female vs 44 male intervention groups) and the missing data for the time of day (22 out of 48), these two moderators were not considered realistic representations and thus were not tested in the moderator analysis.

#### 3.4.2 Moderator analysis (categorical)

##### Training status

Samples experienced (k=28, g=0.38, 95%CI 0.16; 0.60) and inexperienced with resistance training (k=20, g=0.39, 95%CI 0.19; 0.59) did not differ significantly (Q_M_=0.01, df=1, *p*=.92).

##### Type of exercise

Full body (k=23, g=0.39, 95%CI 0.15; 0.63), lower body (k=20, g=0.45, 95%CI 0.20; 0.69), and upper body exercise trainings (k=5, g=0.17, 95%CI -0.14; 0.48) were not significantly different (Q_M_=3.28, df=2, *p*=.19).

##### Risk of bias

The four categories of the risk of bias assessment, low (k=2, g=0.16, 95%CI -1.36; 1.68), moderate (k=16, g=0.31, 95%CI -0.03; 0.65), serious (k=14, g=0.39, 95%CI 0.12; 0.66), and critical (k=16, g=0.49, 95%CI 0.27; 0.72) risk of bias were not significantly different (Q_M_=4.50, df=3, *p*=.21).

#### 3.4.3 Moderator analysis (continuous)

##### Age

Meta-regression revealed that the age of participants was not a significant moderator of the immediate IL-6 response to exercise (k=48, R²=0%, *p*=.73). It must be considered that the meta-regression was based on mean values of age and did not necessarily represent the sample homogeneously.

##### Exercise volume

Meta-regression revealed that the exercise volume was not a significant moderator of the immediate IL-6 response to exercise (k=48, R²=0%, *p*=.37).

##### Exercise intensity

The exercise intensity was not reported in two studies with three effect sizes (Gadruni *et al*. 2015; Gordon *et al*. 2017). Subsequently, these studies were withdrawn from the analysis. Meta-regression revealed that the exercise intensity was not a significant moderator of the immediate IL-6 response to exercise (k=45, R²=0%, *p*=.37)

##### Exercise dose

The exercise dose was not computed in two studies with three effect sizes (Gadruni *et al*. 2015; Gordon *et al*. 2017). Subsequently, these studies were withdrawn from the analysis. Meta-regression revealed that the exercise dose was not a significant moderator of the immediate IL-6 response to exercise (k=45, R²=0%, *p*=.69)

#### 3.4.4 Moderator analysis (multivariate)

The model considered in the multivariate analysis (including training status, exercise dose, training status * exercise dose, and exercise type) was not significant (coeff. 2:6; F(5, 39)=1.025, *p*=.417, R²=0.83%).

### 3.5 Interleukin-10

#### 3.5.1 Main analysis

The average effect size for IL-10 was small (k=22, g=0.16, *p*=0.19), and its confidence interval was between -0.09 and 0.40, which indicates that in a universe of comparable studies to the one included in this analysis, the mean effect size could fall anywhere in between very small negative and small to moderate positive (Figure 2B). The heterogeneity was large (PI [-0.96; 1.28], tau^2^=0.27), with a large part representing the variance of the true effect (I^2^=75.7%). According to the PI, in the universe of populations represented by the included studies, the true effect in 95% of cases will fall between large negative and large positive effects. No outliers were detected. A potential small visual asymmetry was assumed (Figure 3B) and confirmed by Egger’s test (intercept=-0.89, *p*=.078), but no adjustments were necessary according to the trim-and-fill analysis (SE=2.81).

Due to the skewed distribution in sex (3 female vs 19 male intervention groups) and the missing data for the time of day (10 out of 22), these two moderators were not considered realistic representations and thus were not tested in the moderator analysis.

#### 3.5.2 Moderator analysis (categorical) Training status

Samples experienced (k=13, g=0.09, 95%CI -0.18; 0.36) and inexperienced with resistance training (k=9, g=0.28, 95%CI -0.26; 0.81) did not significantly differ (Q_M_=0.49, df=1, *p*=.48).

##### Type of exercise

As upper body exercise only had one effect (g=0.37, 95%CI -0.05; 0.79), it was removed from the statistical analysis. The results were not significantly different (Q_M_=0.84, df=1, *p*=.36) for full body (k=13, g=0.24, 95%CI -0.14; 0.61) and lower body exercises (k=8, g=0.02, 95%CI -0.37; 0.41).

##### Risk of bias

The four categories of the risk of bias assessment, low (k=2, g=0.12, 95%CI -0.70; 0.93), moderate (k=6, g=-0.05, 95%CI -0.66; 0.57), serious (k=8, g=0.00, 95%CI -0.26; 0.27), and critical (k=6, g=0.64, 95%CI -0.05; 1.33) risk of bias were not significantly different (Q_M_=5.16, df=3, *p*=.16).

#### 3.5.3 Moderator analysis (continuous)

##### Age

Meta-regression revealed that the age of participants was not a significant moderator of the immediate IL-10 response to exercise (k=22, R²=0%, *p*=.81). It must be considered that the meta-regression was based on mean values of age and did not necessarily represent the sample homogeneously.

##### Exercise volume

Meta-regression revealed that the exercise volume was not a significant moderator of the immediate IL-10 response to exercise (k=22, R²=0%, *p*=.72).

##### Exercise intensity

Meta-regression revealed that the exercise intensity was not a significant moderator of the immediate IL-10 response to exercise (k=22, R²=0%, *p*=.51)

##### Exercise dose

Meta-regression revealed that the exercise dose was not a significant moderator of the immediate IL-10 response to exercise (k=22, R²=0%, *p*=.59)

#### 3.5.4 Moderator analysis (multivariate)

The model considered in the multivariate analysis (including training status, exercise dose, training status * exercise dose, and exercise type) was not significant (coeff. 2:6; F(5, 16)=0.517, *p*=.760, R²=0%).

### 3.6 Interleukin-1ra

#### 3.6.1 Main analysis

The average effect size for IL-1ra was large (k =12, g=0.78, *p*<.001), and its confidence interval was between 0.55 and 1.02, which indicates that in a universe of comparable studies to the one included in this analysis, the mean effect size could fall anywhere in between moderate and large (Figure 2C). This range did not include 0, which revealed that the true effect is very likely to be positive and equal or beyond moderate. The heterogeneity was small to moderate (PI [0.14; 1.42], tau^2^=0.08), with a moderate part representing the variance of the true effect (I^2^=44.0%). According to the PI, in the universe of populations represented by the included studies, the true effect in 95% of cases will fall between small and large positive effects. No outliers were detected. Visual asymmetry was assumed and confirmed by Egger’s test (intercept=-1.21, *p*<.010). The trim-and-fill analysis showed two missing studies on the left side (Figure 3C). With these two missing effects imputed, the effect size was reduced but remained significant (*p*<.001), k = 14, g = 0.68 (95% CI: 0.42;0.94).

No studies reported using females only, all included studies were ranked with a critical risk of bias, and no study reported the testing time; thus, the moderator analysis was not performed for sex, risk of bias, and testing time. Additionally, considering the missing information in an already small sample of studies for exercise volume, -intensity, and -type, these potential moderators were not tested as well.

Furthermore, according to the relatively small I^2^, this moderator analysis’ conclusions and extent are limited.

#### 3.6.2 Moderator analysis (categorical)

##### Training status

Samples experienced (k=8, g=0.70, 95%CI 0.36; 1.04) and inexperienced with resistance training (k=4, g=0.97, 95%CI 0.60; 1.34) did not significantly differ (Q_M_=2.09, df=1, *p*=.15).

#### 3.6.3 Moderator analysis (continuous)

##### Age

Meta-regression revealed that the age of participants was not a significant moderator of the immediate IL-1ra response to exercise (k=12, R²=0%, *p*=.81). It must be considered that the meta-regression was based on mean values of age and did not necessarily represent the sample homogeneously.

##### Exercise dose

Meta-regression revealed that the exercise dose was not a significant moderator of the immediate IL-1ra response to exercise (k=12, R²=0%, *p*=.36).

#### 3.6.4 Moderator analysis (multivariate)

The model considered in the multivariate analysis (including training status, exercise dose, and training status * exercise dose) was not significant (coeff. 2:5; F(4, 7)=0.893, *p*=.516, R²=0%).

## 4 Discussion

This systematic review and meta-analysis aimed to examine the effects of acute resistance exercise on the release of the myokines IL-6, IL-10, and IL-1ra. A second aim was to characterize how different exercise parameters may influence this response. To the authors’ knowledge, this paper is the first to statistically evaluate the change in concentration of IL-6, IL-10, and IL-1ra from baseline to immediately post-exercise in relation to different moderators.

### 4.1 Main results

Twenty-six studies were included in this systematic review, of which 24 were considered for the meta-analysis investigating IL-6 (k=48), ten for IL-10 (k=22), and four for IL-1ra (k=12). The main findings were a small to moderate positive mean effect for IL-6 and a large positive mean effect for IL-1ra (slightly decreased after trim-and-fill analysis) immediately post-exercise. No significant effects were detected for IL-10. For all three meta-analyses, the overall heterogeneity was moderate to large, with only a moderate to large part of this heterogeneity representing the variance of the true effect. Asymmetry suggesting eventual small-study effects or publication bias was detected for all three meta-analyses.

#### 4.1.1 IL-6

Only seven out of 48 intervention groups investigating IL-6 documented a negative effect immediately post-exercise, resulting in a significant small to moderate positive average effect. This post-exercise increase in circulating IL-6 levels has also been a consistent finding among studies employing endurance exercise protocols (Kakanis *et al*. 2014; MacDonald *et al*. 2003; Ulven *et al*. 2015; Zaldivar *et al*. 2006). For instance, an investigation of IL-6 kinetics during treadmill running at 75% of maximal oxygen uptake showed that plasma concentrations increased in an almost exponential manner to reach their peak upon exercise cessation (Ostrowski *et al*. 1998).

The mobilization of leukocytes into the bloodstream during exercise, coupled with the heightened transcriptional activity of inflammatory genes in peripheral blood mononuclear cells, indicates that immune cells play a role in the post-exercise elevation of IL-6 (Büttner *et al*. 2007; Connolly *et al*. 2004). IL-6 can stimulate the differentiation of B cells into antibody-producing plasma cells and the growth and differentiation of T cells (Hirano 1998), especially CD4+ T cells, by determining their effector functions (Dienz & Rincon 2009). As IL-6 contributes to the differentiation of T helper cells and the production of further cytokines such as IL-4, IL-21, and IL-17 (Dienz & Rincon 2009), cellular and humoral immunity can be enhanced. In the regenerating muscle, IL-6 is released by infiltrating neutrophils and macrophages (Zhang *et al*. 2013), by fibro-adipogenic progenitors (Joe *et al*. 2010), and by satellite cells (Kami & Senba 1998, 2002), suggesting possible autocrine and paracrine functions of IL-6 in satellite cell-dependent myogenesis (Muñoz-Cánoves *et al*. 2013). However, accumulating evidence from biopsy, gene sequencing, or blood sampling studies (Steensberg *et al*. 2000; Steensberg *et al*. 2002; Toft *et al*. 2011) indicates that IL-6 release from the myocytes of the working muscle is far more important to its systemic rise than by immune cells.

Making a distinction between the cell of origin and the environmental context is of significant importance, as it determines the effect of IL-6. Specifically, the release of IL-6 by immune cells is usually accompanied by IL-1β and TNF-α co-secretion and thus triggers pro-inflammatory signaling cascades and neutrophil infiltration that ultimately mediate tissue repair (Pedersen & Febbraio 2008). Whereas the release of IL-6 by the skeletal muscle is not triggered by immune cell signaling but by augmented calcium signaling, glycogen depletion, and lactic acid accumulation (Kistner *et al*. 2022). The fact that muscle-derived IL-6 is released in response to energetic stress indicates its main tasks: liberating energy, enhancing muscular energy uptake, and transiently dampening immune system activity (Kistner *et al*. 2022). To achieve the latter, muscle-derived IL-6 inhibits the production of TNF-α, while promoting the synthesis of IL-10 and IL-1ra, without activating classical pro-inflammatory pathways (Pedersen 2011b). In addition, as the skeletal muscle also consumes carbohydrates during exercise, several myokines, among others IL-6, promote the expression of glucose transporter 4 in skeletal muscles and increase muscular insulin sensitivity, thus lowering plasma glucose concentrations during exercise and up to 24 hours afterward (Kistner *et al*. 2022; Pedersen & Febbraio 2008). These effects amplify the anti-inflammatory effects to prevent and treat the various forms of diabetes mellitus (Oh *et al*. 2016). Furthermore, IL-6 increases lipolysis in skeletal muscle (Petersen & Pedersen 2005; Wolsk *et al*. 2010) as well as fat oxidation in adipose tissue via activation of 5’ adenosine monophosphate-activated protein kinase (AMPK) (Al-Khalili *et al*. 2006; Carey *et al*. 2006; Kelly *et al*. 2009), reducing adipose tissue with inflammatory capacity. Additionally, it mediates crosstalk between insulin-sensitive tissues, the gut, and pancreatic islets to adapt to changes in insulin demand by increasing glucagon-like peptide-1 secretion (Muñoz-Cánoves *et al*. 2013). In light of this, it can be inferred that acute resistance exercise also facilitates anti-inflammatory and anti-diabetogenic outcomes.

#### 4.1.2 IL-10

Unexpectedly, based on the studies included in the meta-analysis, we could not find a significantly positive average effect for IL-10 immediately post-exercise, indicating that the IL-10 increments elicited by an acute resistance exercise bout might either be less pronounced or delayed compared to the other investigated myokines.

Given the previously mentioned IL-6-stimulated release of IL-10 during exercise, this finding might come as a surprise but may be rooted in the kinetic of its activation. Specifically, it has been demonstrated that, in contrast to IL-6, the primary source of IL-10 in response to exercise are monocytes and lymphocytes (Fischer 2006; Nieman *et al*. 2006). It is therefore conceivable that the employed exercise schemes were not able to induce sufficiently high levels of IL-6 to stimulate significant IL-10 release, potentially due to a short duration or their intermittent character. Drawing a comparison with studies on endurance exercises, it has been observed that prolonged cycling for a minimum of one hour led to notable elevations in IL-10, as reported by Nieman et al. (2006) and Ulven et al. (2015). However, investigations utilizing shorter durations revealed only minor increases in IL-6 and did not show significant changes in IL-10, as demonstrated by Cullen *et al*. (2016) and Markovitch *et al*. (2008). Hence, the authors concluded that the induced IL-6 increase might not have been sufficient to induce downstream systemic anti-inflammatory responses immediately post-exercise (Cullen *et al*. 2016).

Beyond that, it may be important to consider that the kinetic of IL-10 activation through IL-6 has a second branch passing by the kynurenine pathway, potentially requiring more delay. Indeed, the exercise-induced IL-6 increase activates the indoleamine 2,3-dioxygenase and kynurenine 3-monooxygenase enzymes producing more kynurenine and kynurenic acid (Fuchs *et al*. 1990; Joisten, Schumann *et al*. 2020). These metabolites are ligands to the transcription factor aryl hydrocarbon receptor, promoting the differentiation of naïve CD4+ T-cells to regulatory T-cells that are the main producers of anti-inflammatory cytokines, including IL-10 (Joisten, Schumann *et al*. 2020). As muscle contraction-induced IL-6 peaks at the end of the exercise, many studies included in this review also reported a continued IL-10 release between 30- and 60 minutes post-exercise (Agostinete *et al*. 2016; Benini, Prado Nunes, Orsatti, Barcelos, Orsatti 2015; Benini, Prado Nunes, Orsatti, Portari, Orsatti 2015; Gerosa-Neto *et al*. 2016; Lira *et al*. 2020).

In addition to physiological considerations, it has also to be taken into account that methodological features of the included studies might have influenced the results of the meta-analysis. Yet, looking at the two studies with the largest negative effect, no obvious explanation becomes apparent. While Lira et al. (2020), g=-1.00, was assessed to be at moderate risk of bias and did not show any remarkable characteristics for the participants nor the intervention itself, Brenner et al. (1999) neither controlled for the physical activity before exercise nor reported an identical warm-up for all participants which might serve as the best explanation for the divergent results. Still, it is not entirely clear what factors can explain the variability in IL-10 results highlighting the need for further research.

Immunologically, increased levels of IL-10 contribute to an anti-inflammatory environment by inhibiting the synthesis of pro-inflammatory cytokines as well as MHC class II and co-stimulatory molecules in activated macrophages/monocytes (Islam *et al*. 2021). Moreover, IL-10 blocks the release of IL-1α, IL-1β, and TNF-α as well as the production of chemokines, including IL-8 and macrophage inflammatory protein-α from lipopolysaccharide-activated human monocytes (Moore *et al*. 1993; Pretolani 1999). Additionally, it not only inhibits the synthesis of these effectors but also increases the expression of their natural antagonists (Moore *et al*. 2001). IL-10 is also the principal cytokine that suppresses cell-mediated immunity and dendritic cell maturation (Moore *et al*. 2001). For the working muscle, these anti-inflammatory effects of IL-10, and IL-1ra, are of significant importance to ensure energy supply by limiting the energy expenditure of the immune system (Kistner *et al*. 2022). Additionally, IL-10 has been reported to prevent insulin resistance in the muscle (Dagdeviren *et al*. 2017).

#### 4.1.3 IL-1ra

Finally, the results of the meta-analysis revealed a large positive average effect for IL-1ra, supporting our hypothesis that resistance exercise bouts result in acute increases of IL-1ra. The investigated effect was the most pronounced among the myokines investigated, which might be a result of greater homogeneity among included studies. Our results align with previous studies employing other exercise forms showing that running (Scott *et al*. 2011) and cycling (Connolly *et al*. 2004) lead to significant increases in IL-1ra levels and gene expression immediately post-exercise and within one-hour post-exercise, respectively. During and after exercise, IL-1ra is predominantly released by macrophages upon IL-6 stimulation (Fischer 2006; Pedersen & Febbraio 2008). The increased circulating levels become functionally relevant as IL-1ra exerts anti-inflammatory effects by inhibiting IL-1β signal transduction, without inducing any intracellular response (Dinarello 2000; Pedersen & Febbraio 2008).

### 4.2 Moderator analysis

We performed moderator analyses for multiple categorical and continuous moderators to test if they were able to explain the heterogeneity of the effect. However, unexpectedly, none of the moderator analyses (univariate or multivariate) revealed an effect of exercise volume, -intensity, or -dose on changes in IL-6, IL-10, and IL-1ra concentration following resistance exercise.

Pedersen et al. (2007) showed that the magnitude of the post-endurance exercise increase in plasma IL-6 could be explained by the duration and intensity of exercise and the muscle mass involved in mechanical work. However, this assumption could not be confirmed for resistance exercise by our meta-analysis. This might be since IL-6 production during exercise is strongly linked to the local metabolism and signals of metabolic and hormonal stress (Kistner *et al*. 2022). While for endurance exercises, glycogen depletion, lactic acid accumulation, or redox signaling increases with duration and intensity of exercise, for resistance exercise volume might be a more reliable surrogate of metabolic stress, as it is positively correlated with the number of muscle contractions and duration of mechanical work. Specifically, exercising with higher intensities allows fewer repetitions than exercising with lower intensities, thus constituting more of a neuromuscular than a metabolic challenge. For instance, the study conducted by Phillips et al. (2010) comparing low-intensity and high-intensity resistance exercise showed that higher exercise doses lead to greater IL-6 changes. In other words, IL-6 levels increased immediately after exercise for both exercises, but the increase was greater in the low-intensity group. Yet, here again, this assumption cannot be confirmed by our results.

The moderator analysis for IL-10 revealed that neither exercise volume, intensity, nor dose significantly moderated its response to an acute bout of resistance exercise. The literature describes the influence of exercise dose on IL-10 and IL-1ra levels equivocally. For example, it has been suggested that changes in IL-1ra and IL-10 post-exercise are determined by the initial changes elicited in the concentration of IL-6 (Pedersen & Febbraio 2008). In contrast to that, Ihalainen et al. (2014) suggested that changes in IL-1ra concentration depend on the type of resistance exercise and less on IL-6. In a systematic review, Cabral-Santos et al. (2019) investigated the response of IL-10 after acute exercise sessions in healthy adults and could not find an evident relationship between intensity and changes in IL-10 production. However, in contrast to our results, they reported a significant linear correlation between exercise duration and deviations of IL-10 from baseline after an acute resistance exercise session. Nevertheless, since this systematic review also included endurance training studies and did not distinguish between endurance and resistance exercise, it must be expected that their results may not be representative of resistance exercise. In addition, it must be taken into account that most of the studies that investigated endurance training sessions were based on long-distance runs, e.g., marathons (Comassi *et al*. 2015; Krzemiński *et al*. 2016; Nickel *et al*. 2012). Compared to resistance training studies such as those included in this meta-analysis, there is a large discrepancy between the duration of endurance training and the duration of resistance training, which is usually about 60 minutes of intermittent training (Benini, Prado Nunes, Orsatti, Barcelos, Orsatti 2015; Goto *et al*. 2020).

Beyond that, based on the included studies, this meta-analysis showed that the resistance training status could not explain any percentage of the variance between the studies. However, the definition of “resistance exercise experienced” is very broad since participants were considered inexperienced if they were described as untrained or sedentary or did not participate in any kind of regular resistance training six months prior to testing. While the participants in the study by Goto *et al*. (2020) performed resistance training regularly for several years, those investigated by Ihalainen *et al*. (2017) only trained for three months before the testing. Thus, great differences even within this definition can occur, and the result regarding this moderator must be treated cautiously.

The moderator age did not help to explain the differences between the study results for Il-6, IL-10, and IL-1ra. The mean age range (19-47 years) from the studies included in this analysis was relatively constrained and potentially not perfectly representative of each sample. It would be valuable to see whether a wider age range would provide the same results. It would be important, especially for older adults, to find out if their myokine response to resistance exercise is different as optimal muscle aging and optimal metabolic control, both related to the release of muscle-derived IL-6, are indispensable factors in healthy aging (Crescioli 2020).

Concerning biological sex, only a few studies tested the acute effects of an acute bout of resistance training on IL-6, IL-10, and IL-1ra in women only (Benini, Prado Nunes, Orsatti, Barcelos, Orsatti 2015; Luk *et al*. 2021). For example, Benini et al. (2015) tested the myokine concentration changes in response to resistance exercise in men and women simultaneously. They found a significant difference between the male and female intervention groups, with men displaying significantly higher concentration changes immediately post-exercise than women, whereas the female intervention group did not show any significant alteration at all. The authors also hypothesized that differences would occur, as there are sex-specific differences in, for instance, muscular fatigue (Hicks *et al*. 2001) or the inflammatory response following exercise (Gillum *et al*. 2011), and different hormonal profiles in general. However, the small number of studies comparing male and female subjects allow no firm conclusions to be drawn about the influence of biological sex on myokine release. Therefore, it is essential to conduct further investigations exploring the differences between men and women and that future studies avoid mixing results between sexes, as the kinetic of myokine activation might be different.

### 4.3 Limitations and perspectives

Despite important findings, the results of this review and meta-analysis should be interpreted within the context of its limitations. First, while our results assess concentration changes immediately post-exercise, further (meta) research is required to evaluate if an effect dose-response relationship can be displayed when assessing later post-exercise time points (e.g., between 5 min and 3 hours postexercise). However, some studies also collected data at later measurement time points. Comparing the concentration changes of IL-6, IL-10, and IL-1ra, there appears to be altered temporal kinetics between endurance and resistance exercise (Figure 4). While it is clearly stated for endurance exercise that peak IL-6 is reached at the end of exercise or shortly thereafter, followed by a rapid decline to baseline levels (Febbraio & Pedersen 2002; Pedersen 2011a), for resistance exercise the time course seems to be much more inconsistent. Moreover, since the exercise-induced production of IL-10 is stimulated by IL-6 (Walsh *et al*. 2011), it could have been assumed that the time course of IL-10 follows the one from IL-6. Unexpectedly, the concentration of IL-10 shows almost a U-shaped curve with great changes directly after exercise and 45 to 60 minutes post-exercise. Islam *et al*. (2021) pointed out that, especially when taking low to moderate-intensity exercise into account, small to no differences were observed in IL-6 concentration compared to non-exercising control groups, whereas a significant change in IL-10 could be observed (Morettini *et al*. 2015; Wadley *et al*. 2016). Therefore, due to these inconsistencies in IL-6 but consistent findings for IL-10, it might be questionable whether this relationship between IL-6 and IL-10 is causal (Islam *et al*. 2021) or if additional factors have a greater influence on IL-10 concentration. Regarding IL-1ra, no firm conclusions can be drawn about changes over time after exercise cessation as no significant data is available, which is not surprising as it is an acute-phase protein (Petersen & Pedersen 2005). Based on these preliminary results, especially high-quality studies on which meta-research can be based is needed to provide further insides into the changes in myokine concentrations after endurance as well as resistance exercise over time.

**Figure 4.**
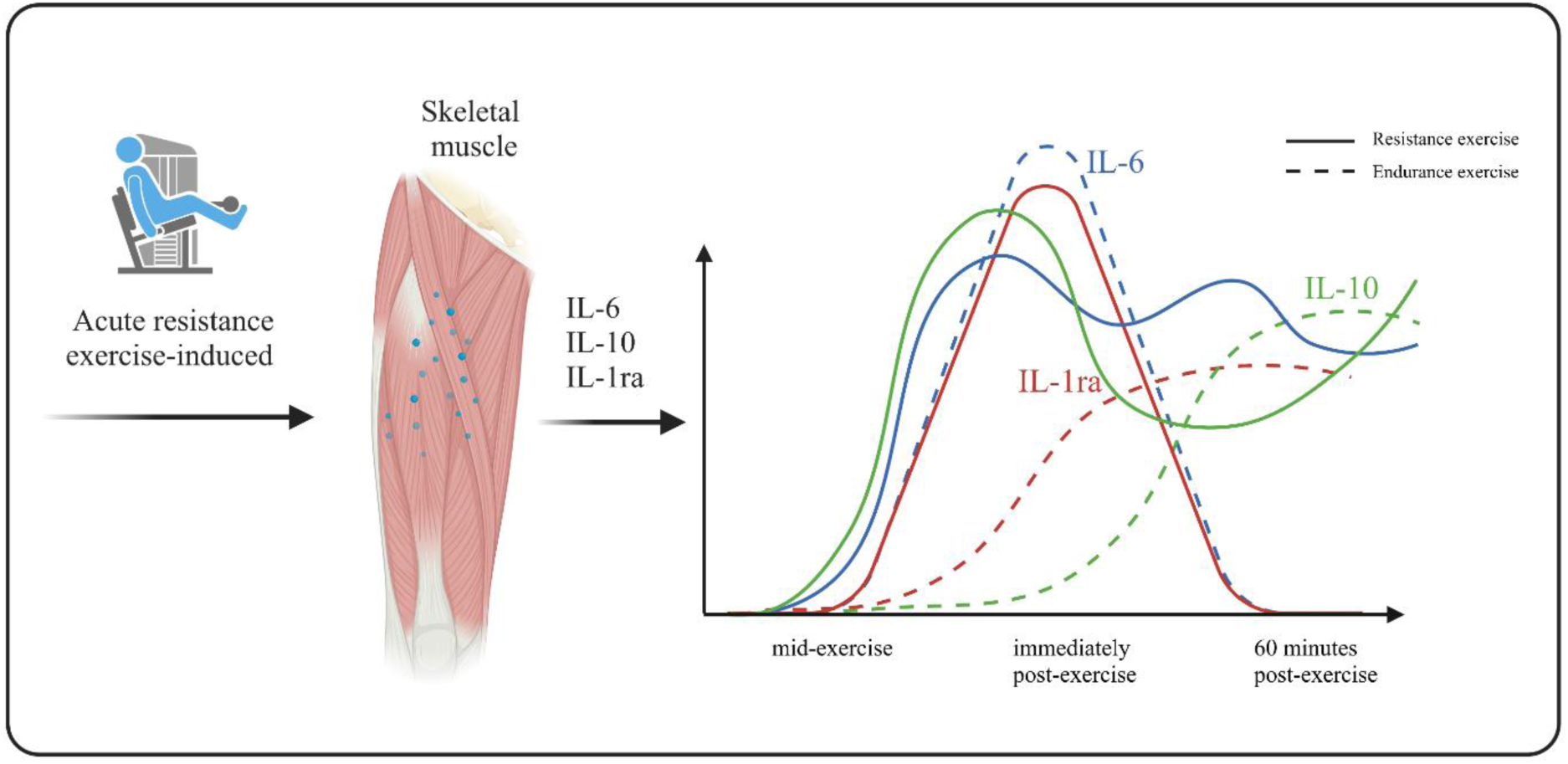
Schematic overview of the changes in concentration of IL-6, IL-10, and IL-1ra for endurance exercise (Pedersen & Febbraio 2008) depicted in comparison to resistance exercises based on the results of the current meta-analysis (created with BioRender.com). mid-exercise: ***, immediately post-exercise: 0-5 min post-exercise.

Second, in the current review, we used exercise volume and intensity to compute the resistance exercise dose. Yet, other training parameters might also be highly relevant. For example, inter- and intra-set rest or proportion of concentric and eccentric loading could be considered. However, most studies did not capture these parameters to a sufficient extent to use them for quantitative analyses, which increases the need for future studies to investigate the different parameters that influence the dose of resistance training. Third, sixty-two percent of the studies were deemed to be at either critical or serious risk of bias, and only one study was of very good quality. The most problematic domain was the confounding variables since most were not controlled for or reported. For instance, the time of the day at which testing was executed was frequently not reported, impairing the possibility of evaluating its effect on the results. Pledge et al. (2011) were the only ones looking into the difference between a morning and evening resistance training session. They were not able to find a significant difference between the exercising groups. However, the control group displayed significantly different baseline values depending on the time point, which may lead to the assumption that IL-6 concentration fluctuates over the day. Further studies should report this more systematically, as one can assume that it might partially influence the results. This is also true for other confounding variables such as diet or the pre-intervention physical activity, including an identical warm-up for all participants. As the studies aimed to determine the effect of resistance exercise on specific myokines, it is indispensable to control for physical activity performed before testing. Indeed, as physical activity can induce, for example, small inflammatory reactions due to minor muscle damage (Severinsen & Pedersen 2020), it can be assumed that they can also affect the study results if they are not controlled and properly reported. Since the diet also affects the immunological profile and possibly leads to an increase in some inflammatory biomarkers (Galland 2010), food intake must be recorded very accurately before testing and should be, at best, standardized for all participants. Another recurrent problem was the small sample size per study. It did not only lead to a large variance per study but also raised questions concerning the representativity of the sample. Finally, some authors also overestimated their effect sizes using the between-subject formula without adjusting it to their within-subject design. This may partly explain why, while the moderator analysis for risk of bias did not show significant differences, the effects in our sample were consistently greater in the three meta-analyses for the studies with high risk of bias. Overall, to improve the general quality of the studies and their representativeness, it would be necessary to provide more information about confounding variables, such as the time of the day. Moreover, variables such as diet, identical warm-up, or pre-intervention physical activity were not controlled as well as they should have been, and therefore, the risk of bias is increased further. Consequently, future research should focus on precisely describing and controlling these influencing factors, as they may change the results to a large extent. In addition, studies with larger sample sizes are needed to avoid overestimating effect sizes affecting the significance of the results.

## 5 Conclusion

The results highlighted in this systematic review and meta-analysis showed a small to moderate positive effect of resistance exercise on IL-6 and a large positive effect on IL-1ra. This could, however, not be shown for IL-10, potentially due to a large sampling variance and a different kinetic of activation. In general, more data on all myokines, especially on IL-10 and IL-1ra, is needed concerning training volume and intensity with more consistency in reporting training parameters before and during the training (e.g., reporting the testing time, controlling physical activity and nutritional intake before the testing session and bigger sample sizes, testing males and females independently), as no clear conclusion on the moderators, in particular, the dose-response relationship can be drawn.

## Data Availability

All data produced are available online at https://osf.io/5jkfa/.

## 6 Conflict of Interest

The authors declare that the research was conducted in the absence of any commercial or financial relationships that could be construed as a potential conflict of interest.

## 7 Author contributions

MR, FJ, SH, and CP designed and wrote the paper. MR, FJ, SH, WB, LF, SB, SD, PHR, MWP, HW, HHWG, and CP were involved in the analysis or interpretation of data and contributed to draft the manuscript or revised it critically for important intellectual content and approved the submitted version.

## 8 Funding

This study is part of the research project Resistance Training in Youth Athletes that was funded by the German Federal Institute of Sport Science (ZMVI4-081901/20-23).

## 9 Acknowledgements

We thank Andreas Stallmach for his important contribution to the discussion of this manuscript.

## 11 Appendix

Supplementary material 1

PRISMA checklist (see external file)

## Notes

### Competing Interest Statement

The authors have declared no competing interest.

### Summary of Updates

The overall layout of the manuscript has been adapted as well as the contributing authors, abstract, section 7, 8, and 9.

